# Community education through local spheres of influence and lived experience of health benefits improve population adherence to programmatic Mass Drug Administration in a persistent schistosomiasis hotspot: an ethnographic study

**DOI:** 10.1101/2024.03.01.24302915

**Authors:** Odoi Paskari, Stella Neema, Birgitte J. Vennervald, Edridah M. Tukahebwa, Shona Wilson

## Abstract

**Background:** The WHO Neglected Tropical Disease Roadmap update for 2021–2030 includes new goals of elimination of schistosomiasis as a public health problem in all endemic countries. Despite heightened efforts since 2012, critical action is still required in addressing barriers to Mass Drug Administration, the primary method of control. This includes improvement in adherence by the populations in persistent schistosomiasis hotspots. One such hotspot is the shoreline of Lake Albert, Uganda, where schistosomiasis control is provided to school-aged children and adults. An overemphasis on regular treatment, without comprehensively addressing factors that result in low uptake of treatment in these high-risk populations is likely to impact the elimination of schistosomiasis as a public health problem.

**Methods:** An ethnographic study using in-depth interviews, key informant interviews, focus group discussions and participant observation was conducted in two study sites along Lake Albert. Thematic content analysis was used during data analysis.

**Results:** The study revealed that the size, taste and smell of the drug, along with its side-effects; poor community integration and occupational behaviour resulting in non-mobilisation; and unfounded rumours and beliefs remain reasons for persistent low uptake of praziquantel by some. Conversely, lived experience of improved health through participation and knowledge of the dangers of the disease if not treated, facilitated treatment uptake. Social influence in crucial knowledge attainment was clear through positive attitudes to localised sensitisation by community drug distributors, along with the delivery of the drug at no cost at home. Crucially, for the majority of participants the facilitating factors were found to outweigh the inhibitory factors related to the drug’s side effects.

**Conclusion:** We recommend a good community engagement strategy that provides continuous education and sensitisation, with improved recruitment and training provision for Community Drug Distributors to facilitate programme reach to groups with current poor engagement.

**Author summary:** Over the last two decades, in the Lake Albert region, Uganda, there has been a number of interventions targeted at schistosomiasis by grass root structures, and district and national level actors; but despite this the Lake Albertine districts remain a highly endemic region for schistosomiasis. In recognition of this persistent schistosomiasis, we examined the factors that inhibit or facilitate adherence to mass drug administration (MDA) using an ethnographic approach. Lived experience of improved health through participation and knowledge of the dangers of the disease if not treated, facilitated treatment uptake. Localised social influence was crucial in gaining knowledge that facilitated uptake. Key were positive attitudes to sensitisation conducted by community drug distributors; whilst uptake of treatment by children was reportedly heavily influenced by their mothers’ positive attitudes to treatment. The drug itself, praziquantel, was described as “very strong” and “effective” because of the relief from symptoms. There are, however, a number of people, both children and adults, who fail to adhere to MDA. Therefore, we recommend continuous education and sensitisation, alongside increased number and training of Community drug distributors/village health team members; with continued motivation for them as they are vital in enabling treatment uptake.

## Introduction

Schistosomiasis, caused by trematode worms of the genus *Schistosoma*, is one of the neglected tropical diseases (NTDs); a group of infections that disproportionately affect low income populations. It remains a major public health problem in tropical and subtropical regions with an estimated 220 million people infected (1)(2). Transmission occurs through contact with water that has been contaminated with faeces or urine containing parasite eggs, and in which intermediate snail hosts breed. The urogenital or gastrointestinal tracts may be impacted depending on the infecting species. Infection can be associated with non-specific morbidities such as anaemia, growth retardation, impairment of cognitive development and work capacity; whilst long-term exposure to infection is associated with specific severe morbidities that can be fatal (3). In Sub-Saharan Africa, schistosomiasis is common, particularly in low - income areas without access to clean water and proper sanitation. At least 90% of those in need of schistosomiasis treatment are living in Africa (4).

Interventions to control schistosomiasis have varied over time and space. Early national control programmes, such as the Sudanese Blue Nile Health Project (started in 1979) and the Egyptian national programme (started in 1988), used a combination of chemotherapy, snail control, provision of clean water and sanitation, and infrastructural development (5, 6). Such multi-approach control programmes are now the exception rather than the rule (7), though the latest edition of the WHO roadmap for control of NTDs emphasises the need to return to multi-approach programmes (8). Instead, due to the combined low cost, good safety record and high efficacy of the anti-schistosomal drug praziquantel (PZQ), preventative chemotherapy (PC) through mass drug administration (MDA) on a population scale became the mainstay of internationally funded national schistosomiasis control programmes. The WHO originally set a 2025 target for these PC programmes of schistosomiasis morbidity control (defined as <5% of school-aged children (SAC) as having a high intensity infection), followed by eradication as a public health burden where possible (8). This has since been revised in the latest Roadmap for NTD control to elimination of schistosomiasis as a public health problem (defined as <1% of school-aged children as having a high intensity infection), in all endemic countries by 2030 (1). However, schistosomiasis elimination efforts have rarely succeeded and only occurred in relatively isolated situations (7). So far elimination has always required extensive (and by extension expensive) integrated approaches, and when not feasible, the tools available have to be optimal to gain the greatest outcomes (9). The elements that result in optimal tools are often not appreciated, hence the need for medical anthropological studies which contribute to our understanding of why programmes are not always successful.

PC interventions involve a variety of elements relating to the target population, the drug to be used and the organisations implementing the intervention. These include population sensitisation, drug procurement and distribution, reporting accuracy and varying involvement of international partners, national government Ministries of Health and Education, and local government and community structures. The interactions of these elements cut across different spheres of influence (from international to local), resulting in non-linear effects on outcomes and a wide range of circumstances may have an impact on the results. In addition, several operational strategies for drug delivery can be adopted depending on the endemicity and national policies. These include variation in platform (school- or community-based), target groups (SAC or SAC alongside adults) and frequency (year or biennially). Given these complexity in PC programme design and implementation, identifying the barriers and facilitators that affect programmes’ ability to meet recommended targets is difficult.

Despite, these complexities, since its internationally supported inception in 2003, MDA using PZQ has led to substantial country-level success in attainment towards the original 2025 goals (10–12). However, regardless of several years of well-implemented MDA coverage, variation in transmission, at times as focal as an individual village, can result in failure to substantially decrease prevalence and mean intensity of schistosomiasis infections at the micro-geographical level (13, 14). Frequently, precision mapping or research studies are required for observation of village-to-village differences. Precise characterisation of these hotspots needs to be applied across the field to understand why some villages fail to decline in their infection status. Only with this understanding can determination of whether enhanced MDA efforts alone will suffice or whether additional control measures will be required in individual areas (14). The new WHO guidelines on treatment now recommend biannually treatment in persistent hotspots (1, 3).

In Uganda MDA began in 2003 through the support of the Schistosomiasis Control Initiative (SCI), and 400,000 treatments of praziquantel were distributed that year (15–17). This has since expanded to the treatment of approximately 1.5 million children annually (18), resulting in significant reductions in *S. mansoni* prevalence, intensity, and associated morbidity (12, 19). Other countries employing MDA have reported similar successes (20). As a result of these findings, WHO’s strategy for disease control in some African nations, including Uganda, has been updated to include the interruption of transmission by 2030 (21). In common with other countries Uganda has areas of continued high schistosomiasis transmission or hotspots; and contributing to this continued high transmission could be coverage rates of the programme in these areas. Social factors are likely amongst the factors that play an important role in MDA uptake. A case in point, an individual’s positioning in the community’s social network, trust in the community medicine distributors (CDDs) for health advice (22), and engagement by the community leaders (23) have all been noted amongst facilitating factors. Poor leadership, insufficient social engagement, lack of genuine community participation, low motivation by the CDDs and their commitment to schistosomiasis control (23), as well as side effects associated with praziquantel (24, 25) are among the reported inhibiting factors for praziquantel uptake. To be able to avert the inhibiting factors while promoting facilitating factors, there is need to examine the barriers to Mass Drug Administration (MDA) in these persistent schistosomiasis hotspots. The aim of the current paper is to investigate these barriers and facilitators of Mass Drug Administration (MDA).

## Materials and Methods

### Study design

The study employed an applied ethnographic study design with a blend of phenomenology approaches to capture the lived experiences of adults. By lived experience, we mean a representation and understanding of research participant’s human experiences, choices, and options and how those factors influence one’s perception of knowledge. The aim was to capture the whole picture of people’s perceptions on barriers and facilitators of MDA. Two social science trained research assistants, one male and one female, who both spoke the local language and a PhD student, spent six weeks in the study area. Prior to implementation of the study 4-days of training was conducted to reinforce the data collectors’ capacity to collect data using qualitative methods. The training included research ethics, confidentiality and best practices in interviewing techniques and encompassed: 1) an overview of the study protocol, 2) research ethics, including consent taking, 3) a review of tools (Focus Group Discussions (FGD), Key Informant Interviews (KII) and in-depth interviews (IDI) and role plays. The data collectors participated in practical exercises to familiarise themselves with the instrument(s) that they would use in the field and a pre-test was conducted that allowed refining of the tools. The data collection and transcription within the communities was conducted November to December 2019 in Hoima District, Uganda.

### Study setting

The study was conducted in the western sub-counties of Kigorobya, Buseruka and Kabaale, Hoima District, which are located along the shores of Lake Albert. Hoima District is in the mid-western region of Uganda, with Hoima Municipality, the seat of the District Headquarters, around 200 km from Kampala using the direct Kampala – Hoima Road. It shares borders with Masindi and Buliisa Districts in the North, Kyankwazi District in the East, and Ntoroko, Kakumiiro, and Kagadi Districts in the south. To the west, Hoima District stretches to the national border with the Democratic Republic of Congo, located within Lake Albert. The district has a total area of 5735.3km^2^, a land area of 3612.17km^2^, and includes Lake Albert and other water bodies amounting to 2123.13km^2^of water. For occupation, the majority of the people in areas above the Lake Albert basin escarpment (Kigorobya and Buseruka sub-counties) are agriculturalists with emphasis on food crop production, while the majority of those who reside near the shores of lake Albert such as in Kabaale subcounty are fishermen and fisherwomen. However, the recent discovery of oil most especially in Kabaale subcounty has made some people to change occupation to business and industrial work.

### Study participants

The study primarily targeted community members in and around Kaiso and Buhirigi Primary Schools, the participatory schools in the wider FibroScHot project (www.fibroschot.eu), in which this anthropology study is integrated. These study participants mainly speak Alur and Runyoro, the two most frequently spoken local languages in Kaiso and Buhirigi. Other participants included key informants at the district, subcounty, and village levels. Participants were purposively selected. During observations and transect walks in the villages, several people were identified as key informants, often because of their position in the community. Their inclusion in the KIIs and IDIs depended on their interest in the study, schistosomiasis and/or health research and roles in the community. In addition, the fisherfolks were considered leaders of their different small groups, or were named key persons because of their age and duration of stay in the community and involvement in fishing activities. The FGDs participants were identified through assistance from the local council leadership and local FibroScHot Project Coordinators, with the aim of gaining a wide range of perspectives.

### Methods of data collection

#### Key informant Interviews

Key informant interviews were conducted at the district, subcounty, and village levels. These included local government officials and community leaders (civil, social, political, religious, opinion). In total 14 key informants were interviewed.

#### In-depth Interviews

In-depth interviews were conducted with community members (male, female, youths) who had experienced schistosomiasis, in order to understand their perceptions and lived experiences (phenomenology) and coping strategies. A youth may be defined in several contexts. According to the United Nations, a youth is a person aged between 15 and 24 years. According to the African Union, a youth is a person aged between 15 and 35 years. In Uganda, a youth is a person aged 18 to 30 years (26). For the purpose of our study, we utilised the Ugandan definition of youth. A total of 53 in-depth interviews were conducted.

#### Focus group discussions

Focus group discussions were conducted with community members comprising of 6 to 7 participants per group. Group categories included gender and age and were organised separately. A total of 14 FGDs were conducted, in which 97 people participated.

#### Participant observations

Participant Observations were conducted at the landing sites to ascertain the activities people were involved in that would expose them to schistosomiasis infection.

### Data Management

Data was analysed using a summative thematic content analysis approach conducted on a rolling basis from transcription to result extraction. All interviews were recorded digitally, transcribed and typed by the research assistants. Notes collected during the interviews (formal and informal) including participant observation data, were expanded every evening by the data collectors. On return from the field, these notes, mostly in the form of jottings in paraphrases, were expanded into fully developed sentences and paragraphs to make sense of what was written. Recordings from the focus group discussions, in-depth interviews and key informant interviews were transcribed verbatim by the research assistants and quality checked by the principal investigator. The transcripts were entered in Nvivo qualitative analysis package (Version 12) for coding and analysis. After reading through the transcripts to get intimate with the data and to gain understanding of the meaning and overall feeling of the data, similar themes and sub-themes were grouped into codes. The code structure evolved inductively to reflect participants’ experiences and voices. After coding, the categories and themes and their interrelationships were developed by the researchers. The results were categorised and interpreted in relation to the objectives of the study and quotes used extensively to further explain and provide evidence for the emergent themes (27).

### Ethical Considerations

The study protocol was approved Makerere University School of Social Sciences Research Ethics committee (MAKSSREC REF 04.19.283) and Uganda National Council for Science and Technology (UNCST REF No SS 5048). Data collection was conducted while respecting the confidentiality of the study participants. Participants were informed of confidentiality procedures as part of the consent process. Consent forms for each type of instrument were administered before the commencement of the interviews. The consent forms were translated into Alur and Runyoro.

## Results

### Socio-Demographic Characteristics of the Participants

#### In-depth interview

The socio demographic characteristics of in-depth interview participants are shown in Table S1. By age over half of the participants who were interviewed as in-depth interviewees were youths (54.3%); 56.6% were male; 65% were married; just below a half (47%) reported having had some secondary education (Senior 1 to Senior 4), and a further 45% had some primary education; 45 (85%) had previous knowledge about schistosomiasis, and one reported currently having schistosomiasis.

#### Focus group discussion

Of the fourteen FGDs conducted, eight comprised of male participants while six FGDs comprised of female participants. The age of participants ranged from 20-51 years. Most of the participants, especially in Kaiso, were fishermen and fishmongers, whereas in Buhirigi many were engaged in farming and a few in fishing. A few of participants had never been to school, but the majority had at least some primary education, followed by secondary education. Many reported having previously been ill with schistosomiasis, more especially those in Kaiso. The socio-demographic characteristics by FGD are available in Table S2.

#### Key informant interviews

The eleven key informants included district administrative officials and health officials, subcounty chiefs and local councillors.

### Facilitating Factors

#### Participation in Mass Drug Administration (MDA)

Most study participants reported to have been sensitized on the dangers of the disease (schistosomiasis) (‘*bilazia’* by the Alur and ‘e*mpuuka’* by the Banyoro) and on praziquantel (‘*baya*’ as referred to by the Alur) as a drug that is effective in treating schistosomiasis. By living at the lake shores, they knew they were exposed to and at risk of getting the schistosomiasis. They were also aware of the MDA programme and its purpose. This compelled them to participate in MDA as they wanted to get cured and live in good health. One male FGD participant shared that:

> “Some people in the community participated in MDA because they were very much aware that living along the shores of Lake Albert, which is the breeding ground for the organism, puts them at risk of getting schistosomiasis” **(Males 23-30 years, FGD, Kaiso).**

A further identified facilitating factor was that MDA took place within their community. They reported that the medicine was distributed by the people they knew and trusted and that the Community Drug Distributors (CDDs) and Village Health Team (VHTs) brought the medicine to their households, moreover free of charge. Furthermore, on this theme, people participated because they were encouraged and had been registered by the CDDs/VHTs who followed them up in their homes.

> “They also participate because the community drug distributors are people who come from within the community and they know them very well’’ **(KII – Local Leader 1).**

There were those who had seen people who had used the medicine and were healed. They reported that having seen and shared experiences with recovered cases encouraged them to participate. Others participated because they took health issues seriously as their own responsibility. Those who suspected they had been infected, or had symptoms of schistosomiasis, participated as the severity of the disease instigated them to take action.

> “Because some people want to make good use of any opportunity to treat schistosomiasis, if they could be having it” **(IDI – Female, 21-25 years old, Kaiso)**.
>
> “Some participated because they have experienced schistosomiasis and they are aware how terrible it is. So, they want to avoid catching it again” **(IDI – Male, 26-30 years old, Kaiso).**

By gender, more women actively participated than men because they wanted to set a good example to the children and the whole family. They also made sure that their children participate in MDA and took their drugs. For the school going children, the children participated because it was organised in schools and it was perceived to be compulsory.

> “Women were the ones who participated most because they were ever active and eager to participate in everything. Men do not participate because they always claim they have no time – they are ever busy” **(IDI – Male, 41-45 years old, Buhirigi).**

The majority of respondents reported that every member of the community who valued health, and knew the benefits and effectiveness of the medicine, participated in MDA programmes. Specific groups of people were pointed out, like the children who were targeted at school where it was perceived to be compulsory, and those who were supported by their mothers; it was stressed that mothers participated in the MDA as responsible parents.

#### Perceptions of the drug praziquantel

When asked to talk more specifically about praziquantel, the drug for schistosomiasis, all categories of respondents reported that the medicine was “very strong” and “effective”. All those who had previously had schistosomiasis reported that the drug was effective in treating schistosomiasis, providing relief from the symptoms they had suffered. They expressed that they got better, were cured or healed after using it. They also liked that it worked fast.

> “The medicine is very good. When I started taking the medicine, I started improving very fast and the symptoms began to disappear**” (IDI-Male, 31-35 years old, Kaiso).**
>
> “I like its effectiveness. Today I am alive and speaking to you because the medicine helped me” **(IDI-Male, 26-30 years old, Kaiso**).

More of what people liked about the medicine was that it helped or made them discover they had schistosomiasis, as they were not aware of any symptoms prior to treatment, but they were evident through improvement in health on taking the medicine; whilst for others it helped them confirm they did not have symptomatic schistosomiasis.

In probing further about what the participants liked about praziquantel, the theme of local availability and the importance of communication about the disease through CDDS and VHTs that was identified during questioning about the MDA programme more generally, was again apparent. The participants reported they liked the medicine because it was free of charge and was brought to their home. Hence no costs incurred.

> “They like its ability to cure the disease, and moreover it is brought for them at their homes” **(KII-Local Leader 1).**
>
> “They continue taking the drug because their community drug distributors together with the village health team (VHTs) sensitised the community members on the dangers of the disease and the benefits of the drug” **(KII – Local Leader 2).**

There were those who were happy it was available through government services, that they had an opportunity to use. Further still, the service providers were reported to have followed up their patients for adherence, which was appreciated, and they also liked the awareness they got on the dangers of the disease and the benefits of the medicine. This combined with the opportunity they had to share experience with champion patients (i.e., people identified in the community to talk about the effectiveness of treatment) and survivors encouraged them in adherence.

> “They say they like it for the fact that it cures them. Most of the people after taking it tell you they now feel better than before” **(Males 20-35 years, FGD, Buhirigi).**

### Inhibiting Factors

#### Participation in Mass Drug Administration

Poor mobilisation issues were identified during questioning. This was caused by a fishing method called ‘s*alacio*’ which is an Alur concept that refers to a fishing method where fishermen stay in the lake for quite a long time e.g., a month or more. As a result of this fishing behaviour, information may not have reached some people in the community as captured during one of the KII:

> “There are fishermen in this community who spend a long time in the lake fishing. Some of these men can be in the lake for weeks and even a month without returning home” **(KII-Local health worker).**

Ignorance most especially by the Congolese migrants was another reason for non – participation. Ignorance was attributed to the protracted insecurity in Democratic Republic of Congo, resulting in continuous movement of Congolese to the Uganda side of Lake Albert where they settle on the landing sites. Whenever there is relative peace in Congo, there are attempts by some of these migrants to go back to Congo. Thus, the frequent movement of these migrants makes it hard for them to consistently adhere to MDA as captured during one of the IDIs:

> “Some people don’t want to participate in MDA in this community because they are not well informed about schistosomiasis and do not know about the intervention programme” **(IDI – Male, 26-30 years old, Kaiso).**

This finding was common among more peripheral members of the community such as Congolese migrants; contrasting directly for those members of the community who are permanent residents who were often reached out to by the CDDs.

Furthermore, some people did not participate because they knew they were not infected or they thought they were safe and not at risk of getting infected with schistosomiasis. The very old people did not participate because they thought it was useless as they were in their last days of life.

> “Some people are just defiant. Some older men especially, claim the disease is not a threat to them because they were born and raised drinking the lake water and they are still healthy” **(KII-Local health worker).**

Perceptions on the impact on fertility was identified as an inhibiting theme, with rumours of reduced sexual desire and some men reported not to having participated because they believed the drug made them infertile or made them impotent. Pregnant women were reported not to have participated because they believed the drug to cause abortion or were informed that it would affect them in some other way and recommended not to participate:

> “Some people don’t want to take the medicine because of rumours that medicine was brought to reduce their sexual desire to control reproductive rate. This is common belief among some people here in this community” **(Males, 31-74 years, FGD, Kaiso).**
>
> “It is because some women are pregnant so they are not supposed to take the medicine Prazinquantel” **(IDI - Female, 26-30 years old, Kaiso).**

Wider non-programme specific social factors were also found to have an impact. It was reported that political influence was one reason for non-participation with members of the opposition discouraging people to take the drug.

> “There is also political influence from opposition who persuade people not to take the drug saying that it is bad, with a fake aim of decampaigning and frustrating government programmes for their own political gain” **(KII - District official).**

Other people did not participate due to religious affiliations and beliefs. These were religions which do not allow use of any medicines, except prayers. One of such group was the Triple six (666) who don’t take medicines as stated by one the IDI participant:

> “Here we have the religious sect called Triple Six (666). They don’t take the medicine because their religion does not allow them to take medicine of any kind” **(IDI-Male, 26-30 years old, Kaiso).**

Finally, other people were said to be careless with their lives, including drunken men who did not care about their lives, and hence did not participate.

> “Others don’t take because of ignorance and carelessness as you can see when you move through the village, there are some categories of people who generally don’t care about life” **(KII-District official).**

#### Perceptions of the drug praziquantel

When asked about factors that may inhibit participation in MDA, the majority of the respondents reported that people feared the side effects of the drug (praziquantel). They reported side effects that included diarrhoea, dizziness and nausea amongst others. The commonest side effects presented were stomach pain/ache and diarrhoea. Other side effects included vomiting, body weakness, body rashes, dizziness, swelling of the body particularly the stomach, giving a bad odour/smell, loss of appetite, serious headache, itching of the body, change in body/skin and hair colour and texture. These side effects were reported to eventually clear with time. Further, the big size, taste and smell of the medicine were other reasons for non-participation. However, it was found that when the participants had lived experience of improved health after taking the drug, the side effects were not inhibitory:

> “People say the medicine is very strong and effective and one should take it when he/she is well prepared for it physically by eating enough and psychologically because of the side effects” **(KII – Local health worker).**
>
> “I have personally taken this medicine. It is good and it helped me, but it made me pass a lot of diarrhoea and I became weak” **(Males, 22-35yrs, FGD, Buhirigi).**
>
> “Praziquantel causes stomach ache, diarrhoea and vomiting, but that is how it eliminates the germs from the body” **(IDI - Female, 26-30 years old, Kaiso).**

There were also some misconceptions. The drug was reported to have treated other ailments by killing other infections. It was also reported to have been liked because it prevented further schistosome infections.

### Motivation to continue participation

A number of factors were reported to motivate people to continue with taking the drug in the future. First, there was desire for complete healing. Some people had hoped they would get well because they were seeing some improvement after starting on the treatment. Some in-depth interview participants reported they wanted to continue with the drug because they were aware of the continued risks of disease if they didn’t take the drug when still exposed to lake waters; and thus, exposure to the disease. Hence, they continued taking it to prevent disease. Motivation was enhanced because they had seen and heard of the survivors’ testimonies. They also knew the dangers of the disease, that if not treated it could be fatal and they knew it had killed people in their community.

> “I continued taking the medicine because I found it of great help to my situation and also, I feared to contract the disease again because I still live in the same landing site with the threat of the disease” **(IDI - Male, 31-35 years old, Kaiso).**

### Why some people did not want to take the medicine

Despite the motivation indicated above, various reasons cut across all categories of respondents for some individuals not wanting to take the medicine. Largely, these can be attributed to the drug itself. Firstly, the fear of the side effects caused by the drug. Secondly, the majority complained about the size of the tablet, as it was too big to be swallowed. Thirdly, the bad odour (smell) and taste were deterrents. The reported negative attitudes/myths towards taking the drugs such as causing impotence and controlling births were also cited as a reason for not wanting to participate. However, despite these myths and fears, most participants reported they were willing and had nothing to prevent them from taking their medicines.

> “Nothing can prevent me from taking the medicine because the benefits of the medicine are more than the side effects” **(IDI - Male, 26-30 years old, Kaiso).**
>
> “Following the threat the disease gave to my life, I don’t think anything can prevent me from taking the medicine” **(IDI - Male, 61-65 years old, Kaiso).**

## Discussion

The three high *S*. *mansoni* endemicity sub-counties of Buseruka, Kabaale and Kigorobya, are situated in Hoima District, Western Uganda which has a long Lake Albert shoreline. This geography, combined with the socio-ecological interactions between the lake shore communities and the lake itself, results in this area being one of the most significant hotspots of schistosomiasis transmission globally. This is despite concerted effort over the last two decades of a preventative chemotherapy (PC) control programme, delivered via the mechanism of MDA. To determine the influence of social factors in participation in the control programme, this study examined the barriers and facilitators of MDA within these communities.

As for the facilitating factors, most study participants reported that they had been sensitised on the dangers of the disease (schistosomiasis) and on praziquantel as a drug that is effective in treating schistosomiasis and they were aware of the MDA programme and its purpose. This compelled them to participate in MDA because they wanted to get cured and live in good health. We additionally found that those who suspected having been infected, or those with symptoms of schistosomiasis participated in treatment to a higher degree. Additionally, participants knew that receiving no treatment can lead to death, therefore such fears compelled them to participate. They reported that they wanted to continue with the drug because they were aware of the risks they were exposed to if they did not take the drug when still exposed to lake waters and by extension to the disease; thus, the preventative rather than curative benefits of participation were also understood. Several studies from elsewhere in Uganda also reveal that the lived experience of the health benefits of treatment, and of knowing others who have had improved health as result of taking praziquantel, leads to MDA uptake (24, 28, 29); while quantitative analysis has also shown that knowledge of the disease is a positive predictor of treatment (30). Systematic review analysis also reveals that people participate in PC programmes as a result of their knowledge and awareness of the signs, effects, and transmission cycle of schistosomiasis (3). Together these findings highlight that health communication regarding the disease is vital for strong uptake of PC.

People also participated in MDA programmes because they were encouraged and registered by the CDDs/VHTs who as part of their role took the time to visit them in their homes. The significant role played by CDDs in sensitising the communities is in line with the findings of Corley et al. (31) and Krentel et al. (32) who highlight CDDs as a “valuable human resource” in the motivation and education of communities, promoting the sustainable uptake of NTD activities. Krentel et al went on to recommended the recognition of CDD contribution by programme managers, whilst acknowledging that non-performance in volunteer roles can also inhibit programmatic achievements (32). Fleming et al. (33) also cited positive and negative influences of these volunteers on programmatic outcomes in the Ugandan districts of Kamuli, Pallisa, Mukono, and Yumbe. Similar findings have been reported from the other east African countries of Rwanda (34) and Tanzania (35). Participants further reported that it was not only that the CDDs/VHTs provided information, but that they knew the people who distributed the medicine and placed importance on this. In Mayuge district, Uganda a direct tie (close friendship) between CDDs and community members was associated with enhanced treatment uptake when compared to CDDs without a direct tie (22). This resonates with the findings of our study in Hoima district; emphasising the need for CDDs to be picked from those within the community. The delivery of the drug at home by the CDDs, free of charge, as a facilitator was also a key finding.

When it comes to the inhibiting factors, we found out that the majority of the participants reported that people feared the nasty side effects of praziquantel during MDA. The side effects mentioned include the well documented ones of diarrhoea, dizziness and nausea. Also well documented is their inhibitory effect on the uptake of treatment (30, 36, 37). In the Ugandan context, this has been found both qualitatively (38) and quantitatively (30). Thus, the fear of side effects has been viewed repeatedly as a key inhibiting factor in uptake and adherence to MDA. However, in our study, the benefits of treatment were found to outweigh the fear of these side effects for many participants, and in fact, many linked these side effects to the strength and effectiveness of the drug. The study did not coincide with the months for MDA in the study area, so corroboration of this finding through structured observation of participants reaction to praziquantel treatment was not possible.

Although women are more likely to take part, worries during pregnancy can be an inhibitory factor. Pregnant women were reportedly not participating because they were worried that the drug caused abortion or affected them in some other way, and also reported that they were informed not to participate. WHO has previously indicated that pregnancy is often cited as a reason for non-compliance (39), potentially resulting from the ineligibility of pregnant and lactating women in the early years of the programme; a recommendation which has since been reversed for those beyond the first trimester of preganancy (40). Reproductive and sexual fears amongst men were also reported, through beliefs of the drug making them either infertile or impotent. The same unfounded fear was noted in Mayuge District, Uganda (36) and Morogoro, Tanzania (41). If such misconceptions are not addressed it can drastically limit the uptake of PC.

Our study result has shown that some people were reported to have been ignorant about the programme especially the Congolese migrants. Most of the Congolese migrants in this study were new or recent arrivals in the study area. As a result of protracted insecurity in Democratic Republic of Congo, there is a continuous movement of Congolese to the Uganda side of Lake Albert. Whenever there is relative peace in Congo, there are attempts by some of these migrants to go back to Congo. Thus, the frequent movement of these migrants makes it hard for them to consistently adhere to MDA, encourage education for their children and to develop deep roots in the community. Such lack of or inadequate knowledge among the migrant population has been reported in other studies as highlighted by systematic review (42). Here we acknowledge that the duration spent on data collection (the six weeks of November to December) could have limited data collection from study participants who are subject to this migratory process. Equally important, and related to migration, the study tools were only translated in two local languages of Alur and Lunyoro, and interviews conducted in English, Alur and Lunyoro. Thus, those of other languages, who will be present amongst these lake shore communities, may have been missed amongst our participants as a result of a language barrier.

Poor mobilisation was another reason for non – participation as information may have not reached some people in the community as captured during data collection. This was caused by a fishing method called ‘s*alacio*’ as practiced by some fishermen. In a situation of this nature, Adriko et al. (30) in their study in Mayuge district, Uganda, suggest that improved educational campaigns and effective mobilization could increase health-seeking behaviour and improve community-wide MDA, even to such categories of community members. A study by Coulibaly et al. (43) in Côte d’Ivoire supports these conclusions with findings that anytime was suitable for treatments if people were informed sufficiently in advance.

Some people did not participate because they knew they were not infected, or they thought they were safe and not at risk of getting infected with schistosomiasis. Whilst the very old did not participate because they don’t see the need as they have always lived in the area (and by extension with the disease). This overlaps with research from Zanzibar by Knopp et al. (44) where people reported missing praziquantel because they felt healthy. Therefore, these complex interacting bio-social factors can also contribute to low drug coverages (30), limiting WHO’s aim of eliminating schistosomiasis as a public health problem by 2030 (1).

## Conclusion and Recommendations

Our research findings indicate a range of inhibiting and facilitating factors in the adherence to MDA. The inhibiting factors include, among others, social factors that have a contextual consequence on uptake, such as religious and political influence and positioning within the community that can lead to ignorance of the programme; but also factors directly related to the intervention, notably the side effects, size, taste and smell of the drug. However, facilitating factors such as knowledge of the severity of the disease, and the fact that people take their health seriously have greatly facilitated praziquantel uptake. Equally important, the localised, no cost availability of the drug, combined with sensitisation and drug delivery by known individuals, combined with lived experience, either their own or of others, were perceived to outweigh the negative factors related to the drug and its side effects. Additionally, unlike what has been found in other studies regarding the side effects of praziquantel, there is strong evidence that the participants had a very good understanding about why the side effects occur, their transitory nature and for the majority they are no longer inhibitory to uptake of treatment.

Arising from the study findings we propose that more needs to be done to promote the facilitating factors that have been shown to counter the inhibiting ones, particularly for those who are not receiving the messaging from the CDDs or are worried about unfounded rumours about the drug. The findings highlight a need for continuous education and sensitisation on schistosomiasis causes, prevention and treatment to the community members in the study area, particularly given that most of them are mobile workers with occupations conducted at the lake. A good community engagement plan or strategy is important and should include the use of expert patients/champions/survivors, alongside local leaders and health workers to advocate for the drug and to encourage uptake in the community. Involvement of all stakeholders: district, village, traditional and religious leaders alongside the CDDs and VHTs in mobilising people for participation will be crucial. Sensitisation, recruitment, training and motivation of drug distributors from within the communities is also key. Finally, treatment supporters such as patient’s relatives, community volunteers working with the programme, medical workers, and pharmacists within the community among others could help to assuage fears of side effects and counter unfounded rumours to ensure greater uptake.

## Data Availability

The data that support the findings of this study will be publicly available from UK Data Service ReShare with the identifier(s) [857010]

## Acknowledgments

At the time of drafting this manuscript, the first author Mr. Odoi Paskari was a postgraduate student in Medical Anthropology at the Department of Sociology and Anthropology, School of Social Sciences, Makerere University. We are grateful for the support from the Department, as well as the implementation research team under the FibroScHot project. The authors would also like to acknowledge the contribution from the Hoima District Health Office and the community members of Kabaale, Buseruka and Kigorobya Sub counties, who were very supportive and ensured that this study was conducted without any significant challenges.

## Financial Disclosure Statement

All authors and study implementation were supported through the FibroScHot project which is part of the EDCTP2 programme supported by the European Union (RIA2017NIM-1842-FibroScHot).

## Competing Interests

The authors have declared no completing interests exist.

## Author Contributions

Conceptualisation (OP, SN, BJV, EMT, SW); data curation (OP, SN); formal analysis (OP, SN); funding acquisition (SN, SW); investigation (OP, SN); methodology (OP); project administration (OP, SN); supervision (SW); validation (OP, SN, SW); writing – original draft (OP, SN); writing – review and editing (SN, BJV, EMT, SW).

## Notes

### Competing Interest Statement

The authors have declared no competing interest.

### Funding Statement

This study was funded by the FibroScHot project which is part of the EDCTP2 programme supported by the European Union (RIA2017NIM-1842-FibroScHot)

### Author Declarations

School of Social Sciences Research Ethics committee of Makerere University gave ethical approval for this work

## References

1. WHO. WHO guideline on control and elimination of human schistosomiasis. World Health Organization. WHO; 2022.

2. WHO. Schistosomiasis: Fact Sheet. WHO; 2019.

3. Torres-Vitolas CA DN, Fleming FM. Factors affecting the uptake of preventive chemotherapy treatment for schistosomiasis in Sub-Saharan Africa: A systematic review. PLoS neglected tropical diseases. 2021;15(1).

4. WHO. Schistosomiasis: Fact Sheet. WHO; 2023.

5. Fenwick A WJ. Schistosomiasis: challenges for control, treatment and drug resistance. Current opinion in infectious diseases 2006;19(6):577–82.

6. el Gaddal AA. The Blue Nile Health Project: a comprehensive approach to the prevention and control of water-associated diseases in irrigated schemes of the Sudan. The Journal of tropical medicine and hygiene. 1985;88(2):47–56.

7. Rollinson D KS, Levitz S, Stothard JR, Tchuenté LA, Garba A, Mohammed KA, Schur N, Person B, Colley DG, Utzinger J. Time to set the agenda for schistosomiasis elimination. Acta tropica. 2013;128(1):423–40.

8. WHO. Accelerating work to overcome the global impact of neglected tropical diseases: a roadmap for implementation. WHO; 2012.

9. Secor WE CD. When should the emphasis on schistosomiasis control move to elimination? Tropical medicine and infectious disease. 2018;3(3):85.

10. Deol AK, Fiona M. Fleming, Beatriz Calvo-Urbano, Martin Walker, Victor Bucumi, Issah Gnandou, Edridah M. Tukahebwa et al. Schistosomiasis — Assessing progress toward the 2020 and 2025 global goals. N Engl J Med, 381. 2019;381:2519–28.

11. Andrade G BD, Gazzinelli A, King CH. Decline in infection-related morbidities following drug-mediated reductions in the intensity of Schistosoma infection: a systematic review and meta-analysis. PLoS neglected tropical diseases. 2017;11(2).

12. Kabatereine NB BS, Koukounari A, Kazibwe F, Tukahebwa EM, Fleming FM, Zhang Y, Webster JP, Stothard JR, Fenwick A. Impact of a national helminth control programme on infection and morbidity in Ugandan schoolchildren. Bulletin of the World Health Organization. 2007;85(2):19–9.

13. Wiegand RE MP, Montgomery SP, Chan YL, Andiego K, Omedo M, Muchiri G, Ogutu MO, Rawago F, Odiere MR, Karanja DM. A persistent hotspot of Schistosoma mansoni infection in a five-year randomized trial of praziquantel preventative chemotherapy strategies. The Journal of infectious diseases. 2017;216(11):1425–33.

14. Kittur N, Sue Binder, Carl H. Campbell Jr, Charles H. King, Safari Kinung’hi, Annette Olsen, Pascal Magnussen, and Daniel G. Colley. Defining persistent hotspots: areas that fail to decrease meaningfully in prevalence after multiple years of mass drug administration with praziquantel for control of schistosomiasis. The American journal of tropical medicine and hygiene. 2017;97(6):1810–7.

15. Fenwick A WJ, Bosque-Oliva E, Blair L, Fleming FM, Zhang Y, Garba A, Stothard JR, Gabrielli AF, Clements AC, Kabatereine NB. The Schistosomiasis Control Initiative (SCI): rationale, development and implementation from 2002–2008. Parasitology. 2009;136(13):1719–30.

16. Fenwick A. New initiatives against Africa’s worms. Transactions of the Royal Society of Tropical Medicine and Hygiene. 2006;100(3):200–7.

17. Clements AC LN, Blair L, Nyandindi U, Kaatano G, Kinung’hi S, Webster JP, Fenwick A, Brooker S. Bayesian spatial analysis and disease mapping: tools to enhance planning and implementation of a schistosomiasis control programme in Tanzania. Tropical medicine & international health. 2006;11(4):490–503.

18. Fleming FM HW, Fenwick A. Schistosomiasis Control Initiative website 2016 [

19. French MD CT, Gambhir M, Fenwick A, Webster JP, Kabatereine NB, Basáñez MG. Observed reductions in Schistosoma mansoni transmission from large-scale administration of praziquantel in Uganda: a mathematical modelling study. PLoS neglected tropical diseases. 2010;4(11).

20. Ouedraogo H DF, Zongo D, Bagayan M, Bamba I, Pima T, Yago-Wienne F, Toubali E, Zhang Y. Schistosomiasis in school-age children in Burkina Faso after a decade of preventive chemotherapy. Bulletin of the World Health Organization. 2016;94(1).

21. WHO. World Health Organization. (2020). Summary of global update on implementation of preventive chemotherapy against neglected tropical diseases in 2019. Weekly epidemiological record. 2020;39(95):469–74.

22. Chami GF KA, Bulte E, Fenwick A, Kabatereine NB, Tukahebwa EM, Dunne DW. Community-directed mass drug administration is undermined by status seeking in friendship networks and inadequate trust in health advice networks. Social science & medicine. 2017;183:37–47.

23. Mwanga JR KhS, Mosha J, Angelo T, Maganga J, Campbell Jr CH. Village response to mass drug administration for schistosomiasis in Mwanza region, northwestern Tanzania: are we missing socioeconomic, cultural, and political dimensions? The American Journal of Tropical Medicine and Hygiene. 2020;103(5):1969.

24. Parker M AT, Hastings J. Resisting control of neglected tropical diseases: dilemmas in the mass treatment of schistosomiasis and soil-transmitted helminths in north-west Uganda. Journal of biosocial science. 2008;40(2):161–81.

25 . Kabatereine NB FF, Nyandindi U, Mwanza JC, Blair L. The control of schistosomiasis and soil-transmitted helminths in East Africa. TRENDS in Parasitology. 2006;22(7):332–9.

26. Statistics UBo. The national population and housing census 2014-main report. Kampala 2016.

27. Creswell JW CJ. Research design: Qualitative, quantitative, and mixed methods approaches: Sage publications; 2017.

28. Fleming FM FA, Tukahebwa EM, Lubanga RG, Namwangye H, Zaramba S, Kabatereine NB. Process evaluation of schistosomiasis control in Uganda, 2003 to 2006: perceptions, attitudes and constraints of a national programme. Parasitology. 2009;136(13):1759–69.

29. Parker M AT. Does mass drug administration for the integrated treatment of neglected tropical diseases really work? Assessing evidence for the control of schistosomiasis and soil-transmitted helminths in Uganda. Health research policy and systems. 2011;9(1):1–20.

30. Adriko M FC, Carruthers LV, Moses A, Tukahebwa EM, Lamberton PH. Low praziquantel treatment coverage for Schistosoma mansoni in Mayuge District, Uganda, due to the absence of treatment opportunities, rather than systematic non-compliance. Tropical medicine and infectious disease. 2018;3(4):111.

31. Corley AG TC, Glass NE. The role of nurses and community health workers in confronting neglected tropical diseases in sub-Saharan Africa: a systematic review. PLoS neglected tropical diseases. 2016;10(9).

32. Krentel A GM, Mallya S, Boadu NY, Amuyunzu-Nyamongo M, Stephens M, McFarland DA. Review of the factors influencing the motivation of community drug distributors towards the control and elimination of neglected tropical diseases (NTDs). PLoS neglected tropical diseases. 2017;11(12).

33. Fleming FM MF, Hansen KS, Webster JP. A mixed methods approach to evaluating community drug distributor performance in the control of neglected tropical diseases. Parasites & vectors. 2016:1–5.

34. Basinga P GP, Binagwaho A, Soucat AL, Sturdy J, Vermeersch CM. Effect on maternal and child health services in Rwanda of payment to primary health-care providers for performance: an impact evaluation. The Lancet. 2011;377(9775):1421-8.

35. Leonard KL MM. Professionalism and the know_Jdo gap: Exploring intrinsic motivation among health workers in Tanzania. Health economics. 2010;19(12):1461–77.

36. Mujumbusi L NE, Ssali A, Pickering L, Seeley J, Meginnis K, Lamberton PH. Understanding perceptions of schistosomiasis and its control among highly endemic lakeshore communities in Mayuge, Uganda. PLoS Neglected Tropical Diseases. 2023;17(1).

37. Burnim M IJ, King CH. Systematic review of community-based, school-based, and combined delivery modes for reaching school-aged children in mass drug administration programs for schistosomiasis. PLoS neglected tropical diseases. 2017;11(10).

38. Kabatereine N FF, Thuo W, Tinkitina B, Tukahebwa EM, Fenwick A. Community perceptions, attitude, practices and treatment seeking behaviour for schistosomiasis in L. Victoria islands in Uganda. BMC research notes. 2014;7(1):1-.

39. WHO. Crossing the Billion. Preventive chemotherapy for neglected tropical diseases: Lymphatic filariasis, onchocerciasis, schistosomiasis, soil-transmitted helminthiases and trachoma. World Health Organization; 2017.

40. WHO. World Health Organization. Preventive chemotherapy in human helminthiasis. Coordinated use of anthelminthic drugs in control interventions: a manual for health professionals and programme managers.: WHO; 2006.

41. Hasting J. Rumours, riots and the rejection of mass drug administration for the treatment of schistosomiasis in Morogoro, Tanzania. Journal of biosocial science. 2016;48(S1):S16–39.

42. Sacolo H CM, Kalinda C. Knowledge, attitudes and practices on Schistosomiasis in sub-Saharan Africa: a systematic review. BMC infectious diseases. 2018;18:1–7.

43. Coulibaly JT OM, Barda B, Utzinger J, N’Goran EK, Keiser J. A rapid appraisal of factors influencing praziquantel treatment compliance in two communities endemic for schistosomiasis in Côte d’Ivoire. Tropical medicine and infectious disease. 2018;3(2):69.

44. Knopp S PB, Ame SM, Ali SM, Muhsin J, Juma S, Khamis IS, Rabone M, Blair L, Fenwick A, Mohammed KA. Praziquantel coverage in schools and communities targeted for the elimination of urogenital schistosomiasis in Zanzibar: a cross-sectional survey. Parasites & vectors. 2016:1–4.

