## Supplementary tables 1 and 2 for "Community education through local spheres of influence and lived experience of health benefits improve population adherence to programmatic Mass Drug Administration in a persistent schistosomiasis hotspot: an ethnographic study"

**Supplementary Table 1**: **Socio-demographic characteristics of the in-depth interview participants**

| **Age range in years** | **N (%)** |
| --- | --- |
| 18-30 | 30 (54.3%) |
| 31-40 | 16 (26.0%) |
| 41-50 | 6 (13.0%) |
| 51+ | 3 (6.5) |
| **Sex** |  |
| Male | 30 (56.6%) |
| Female | 23 (43.3%) |
| **Education Level** |  |
| None | 1 (1.8) |
| Primary | 24 (45.2%) |
| Secondary | 25 (47.1%) |
| Tertiary | 3 (5.6%) |
| **Marital Status** |  |
| Single | 15 (26%) |
| Married | 33 (65.2%) |
| Separated | 4 (6.5%) |
| Widowed | 1 (2.1%) |
| **Reported *S. mansoni* Infection Status** |  |
| Ever had | 45 (84.9%) |
| Currently have | 1 (1.8%) |
| Never had | 7 (13.2%) |

**Supplementary Table 2:** **Socio-demographic Characteristics of the FGD participants**

| **FGD** | **Site** | **Number of participants** | **Age Range/Category** | **Final Year Education** | **Gender** |
| --- | --- | --- | --- | --- | --- |
| 1 | Kaiso | 6 | 25-42 | P3 -S1 | Female |
| 2 | Kaiso | 7 | 20-24 | P7-S4 | Male |
| 3 | Kaiso | 7 | 19-38 | P3-S1 | Female |
| 4 | Kaiso | 7 | 38-57 | 0-P6 | Female |
| 5 | Buhirigi | 7 | Youths | P4-S3 | Male |
| 6 | Buhirigi | 7 | Youths | 0-S4 | Female |
| 7 | Buhirigi | 7 | 33-50 | 0-P6 | Female |
| 8 | Buhirigi | 7 | 20-41 | P2-S4 | Male |
| 9 | Kaiso | 7 | 23-30 | P5-S4 | Male |
| 10 | Kaiso | 7 | 31-74 | P5-S4 | Male |
| 11 | Buhirigi | 7 | 23-35 | P1-S2 | Male |
| 12 | Buhirigi | 7 | Youths | P4-S2 | Male |
| 13 | Buhirigi | 7 | 36-51 | P4-S2 | Male |
| 14 | Buhirigi | 7 | 37-58 | 0-P7 | Female |
